# IFNL4 genetic variant can predispose to COVID-19

**DOI:** 10.1101/2021.03.01.21252696

**Authors:** Jose Maria R Saponi-Cortes, Maria Dolores Rivas, Fernando Calle, Juan F Sanchez Muñoz-Torrero, Alberto Costo, Carlos Martin, Jose Zamorano

## Abstract

Interferon lambda 4 (IFNλ4) has shown antiviral activity against RNA viruses, including some coronaviruses. Besides, genetic variants of IFNL4 can be predictive of clearance of RNA viruses. However, little is known about the effect of these genetic variants on SARS-CoV-2 infection. In this study, we investigated whether there was a relationship of the rs12979860 polymorphism of IFNL4 with COVID-19. We found that the T allele of rs12979860 was overrepresented in the COVID-19 patients with regard to the general population without this disease (36.16% vs. 26.40%, p=6.4×10^−4^; OR=0.633; 95% CI=0.487,0.824), suggesting this allele could be a risk factor to COVID-19. Accordingly, the CC genotype was significantly lower in COVID-19 patients compared to controls (37.85% vs. 55.51%, p=8×10^−5^; OR=0.488; 95% CI=0.342,0.698). These results were not affected by sex, age, and disease severity in patients with COVID-19. Our findings suggest that, like other infectious diseases caused by RNA viruses, genetic variants of IFNL4 may predispose to COVID-19. Confirmation of our results could have a substantial impact on the prognosis and treatment of this disease.

## Introduction

Coronavirus disease-19 (COVID-19) is characterized by a set of symptoms that develop following infection with severe acute respiratory syndrome coronavirus 2 (SARS-CoV- 2) (1). Besides viral genetic variations, it is accepted that social, economic, and host factors can affect the incidence of COVID-19 (2). Thus, the characterization of the host genetic factors contributing to this disease would be of great interest for prognosis and treatment.

Interferons (IFNs) are believed to play an important role in the outcome of COVID-19. This is manifested by the fact that type I IFN deficiencies can lead to severe disease (3). However, trials investigating the efficacy of type I IFN treatments have shown inconsistent results (4,5). Growing evidence suggests that type III IFNs may also play a role in host defense against SARS-CoV-2 (6). Among them, little is known about the role of interferon lambda 4 (IFNλ4), even though it has shown antiviral activity against RNA viruses, including some coronaviruses (7). The importance of this cytokine in viral infections was highlighted by the discovery that genetic variants of IFNL4 could be predictive of viral clearance. Thus, the rs12979860 single nucleotide polymorphism has been associated with clearance of hepatitis C virus (HCV) and RNA viruses of the upper respiratory tract and (8-10). Furthermore, this polymorphism is also associated with response to type I IFN treatments in chronic hepatitis C (11). With these antecedents, we investigated whether there was also a relationship of IFNL4 genetic variants with COVID-19. To this end, we analyzed the rs12979860 polymorphism in IFNL4 in a cohort of COVID-19 patients and compared it with the general population without this disease.

## Patients and Methods

This study was conducted to analyze the IFNL4 rs12979860 single nucleotide polymorphism in a population of COVID-19 patients and compare it with a control population without this disease (non-COVID-19). 177 patients diagnosed with COVID- 19 were consecutively recruited for genotyping analysis from April to December 2020 in the post-COVID-19 follow-up at the San Pedro de Alcantara Hospital (Caceres, Spain). The diagnosis of COVID-19 was established based on the presence of symptoms and confirmed by positive SARS-CoV-2 RNA in respiratory secretions and antibodies against SARS-CoV-2 using standard laboratory procedures. When indicated, patients were subdivided into non-severe- (n=132) and severe-COVID-19 (n=45). Severe disease was defined following the criteria of Gandhi et al. as a blood oxygen saturation <94%; respiratory rate ≥30 breaths/min; pulmonary infiltration >50%, requiring oxygen therapy (12). Non-severe COVID-19 includes mild and moderate illness as described by Gandhi et al (12). Non-COVID-19 control samples were obtained from volunteer donors before 2019 to avoid interference from undetected infected donors. Patients and controls were classified by gender, age, and severity of the disease. Table 1 summarizes the population analyzed. The proportion of females and males was similar in both groups. The average age was 68 (±16) years old in the COVID-19 group and 49 (±29) years old in the control group. Individuals equal or over 65 years old represent 58.76% in the patients and 43.6% in the control group. 74.58% of COVID-19 patients suffered non-severe conditions while the rest were classified as severe (25.42%). Patients and controls gave their written informed consent and, when applicable, provided consent for banking and future analysis of biological samples. The study was approved by the Hospital Ethical Committee for Clinical Research.

**Table 1.**
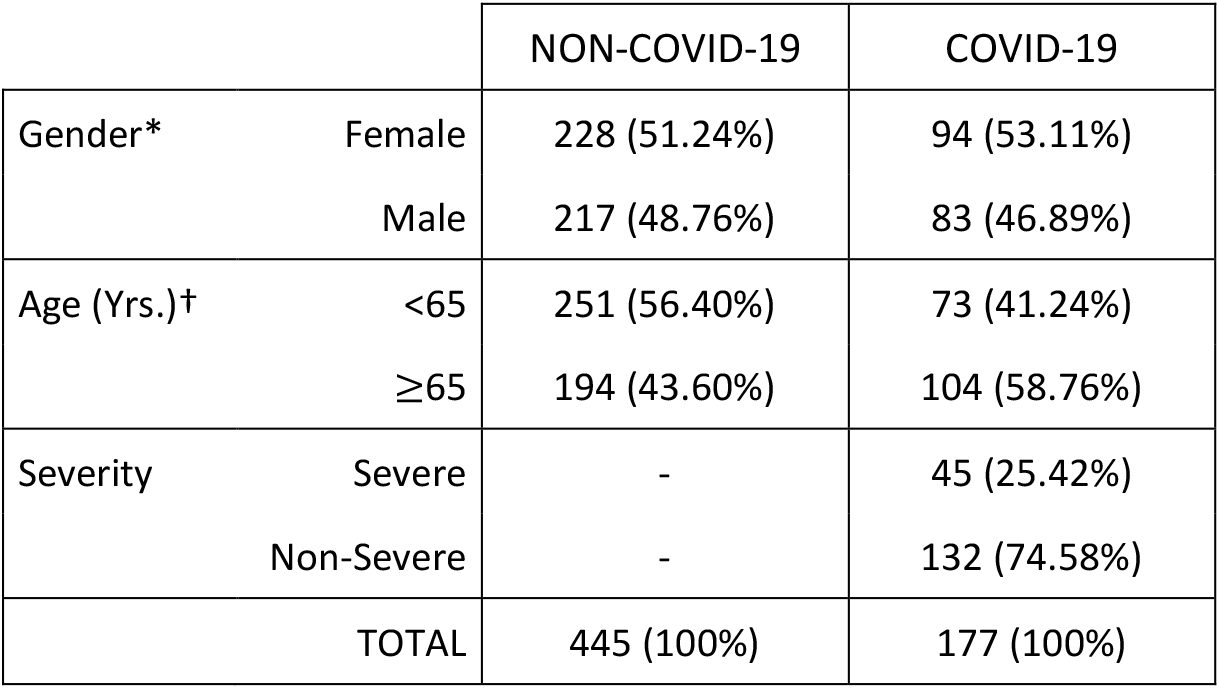
Characteristics of patients and controls. Chi-square test: *p=0.673; OR=1.078; 95% CI=(0.760;1.528). †p=0.01; OR=0.543; 95% CI=(0.381,0.772).

|  |  | NON-COVID-19 | COVID-19 |
| --- | --- | --- | --- |
| Gender* | Female | 228 (51.24%) | 94 (53.11%) |
|  | Male | 217 (48.76%) | 83 (46.89%) |
| Age (Yrs.)† | <65 | 251 (56.40%) | 73 (41.24%) |
|  | ≥65 | 194 (43.60%) | 104 (58.76%) |
| Severity | Severe | - | 45 (25.42%) |
|  | Non-Severe | - | 132 (74.58%) |
| TOTAL |  | 445 (100%) | 177 (100%) |

Genotyping analysis. Genomic DNA was isolated from whole blood using a Nucleospin blood kit (Macherey-Nagel, Duren, Germany). Genotyping to determining rs12979860 alleles was performed in a Stratagene 3005 MxP Quantitative PCR (Agilent Technologies, Inc., Santa Clara, CA) by using a Taqman genotyping mastermix (Thermofisher Scientific, Waltham, MA) and a commercial assay also from Thermofisher Scientific (Assay ID: C__7820464_10).

Statistical analysis. Genotype and allele frequencies were obtained from sample data, comparing between patients and controls marked as COVID-19 and non-COVID-19 respectively. Categorical variables were compared using Pearson Chi-square two-sided test since the frequencies were above 5 in all cases. Normality was assumed since the size of the sample was big enough and z-tests for two proportions were adjusted for all pairwise comparisons within a row of proportions in each variable. Odds ratio (OR) and 95% confidence interval (CI) were calculated when the resulting frequency table was 2×2 (for the dichotomous categorical variables). P-values less than 0.05 were considered statistically significant. Comparison analyses were carried out using statistical software SPSS for Windows Release 25.0 (SPSS, Inc).

## Results

The characteristics of 177 COVID-19 patients and 445 non-COVID-19 controls by gender and age are shown in table 1. The proportion of female and male between COVID-19 patients and controls was similar (p=0.673; OR=1.078; 95% CI=(0.760;1.528). However, the percentage of age ≥65 years was higher in COVID-19 patients (p=0.01; OR=0.543; 95% CI=0.381,0.772), consistent with the fact that older people are the most affected by COVID-19 (1).

The genotype and allele frequencies of the rs12979680 polymorphism are shown in table 2. The T allele was overrepresented in the COVID-19 patients with regard to controls (36.16% vs. 26.40%, p=6.4×10^−4^; OR=0.633; 95% CI=(0.487,0.824), suggesting that this allele could be a risk factor to develop COVID-19. Accordingly, the CC genotype was significantly lower in COVID-19 patients compared to controls (37.85% vs. 55.51%, p=8×10^−5^; OR=0.488; 95% CI=(0.342,0.698). Then, we analyzed whether the ratio of rs12979860 genotypes was affected by sex, age, and severity of disease in the COVID-19 group, comparing the protective CC genotype versus CT+TT (Table 3). Using Chi-square test, we found no significant differences between female and male (p=0.155; OR=0.642; 95% CI=(0.349,1.184), younger and older than 65-year-old (p=0.618; OR=1.145; 95% CI=(0.618,2.119), and severe and non-severe disease (p=0.484; OR=1.279; 95% CI=(0.642,2.549).

**Table 2.** Table of frequencies for rs12979860 genotypes and alleles in patients and controls. Tests are adjusted for all pairwise comparisons by rows using the Z-test.

| IFNL4 rs12979860 | NON-COVID-19 | COVID-19 | p-value | OR (95% CI) |
| --- | --- | --- | --- | --- |
| Genotype CC * | 247 (55.51%) | 67 (37.85%) | $8 \times 10^{-5}$ | |
| CT * | 161 (36.18%) | 92 (51.98%) | $3 \times 10^{-4}$ | |
| TT | 37 (8.31%) | 18 (10.17%) | 0,4593 |  |
| Genotypes CC * | 247 (55.51%) | 67 (37.85%) | $8 \times 10^{-5}$ | 0.488 (0.342, 0.698) |
| CT+TT* | 198 (44.49%) | 110 (62.15%) |  |  |
| Allele C * | 655 (73.60%) | 226 (63.84%) | $6.4 \times 10^{-4}$ | 0.633 (0.487,0.824) |
| T * | 235 (26.40%) | 128 (36.16%) |  |  |

**Table 3.** rs12979860 genotypes by gender, age, and severity rates in COVID-19 patients. P-value from Chi-square test.

|  |  | rs12979860 Genotype |  | p-value | OR (95% CI) |
| --- | --- | --- | --- | --- | --- |
|  |  | CC | CT+TT |  |  |
| Gender | Female | 31 (46.27%) | 63 (57.27%) | 0,155 | 0.642<br>(0.349,1.184) |
|  | Male | 36 (53.73%) | 47 (42.73%) |  |  |
| Age | <65 | 29 (43.28%) | 44 (40.00%) | 0,667 | 1.145<br>(0.618,2.119) |
| | $\geq 65$ | 38 (56.72%) | 66 (60.00%) | | |
| Severity | Severe | 19 (28.36%) | 26 (23.64%) | 0,484 | 1.279<br>(0.642,2.549) |
|  | Non-Severe | 48 (71.64%) | 84 (76.36%) |  |  |

## Discussion

This is a study comparing a genetic variant of IFNL4 in a population of COVID-19 patients with the general population without this disease. Genetic variants of IFNL4 have already been associated with viral clearance of HCV and other RNA viruses (8-11). In this study, we have found that the rs12979860 polymorphism in IFNL4 was also associated with the presence of symptomatic SARS-CoV-2 infection. Thus, the T allele of rs12979860 was significantly overrepresented in COVID-19 patients regarding non-COVID-19 controls, suggesting that this allele may be a risk factor for COVID-19. Accordingly, the frequency of the CC genotype was lower in the COVID-19 group. Our study did not provide evidence to explain this observation; however, the T allele has already been associated with ineffective viral clearance of HCV and other RNA viruses (8-11). In contrast, the CC genotype favors clearance of these viruses (8,10). The rs12979680 polymorphism is in strong linkage disequilibrium with the rs368234815 polymorphism of IFNL4 that determines the production of IFNλ4 (9). Thus, the T allele of rs12979860 is linked to the production of IFNλ4 while the C allele to the inactivation of the IFNL4 gene. Intriguingly, the production of IFNλ4 does not facilitate the clearance of RNA viruses, and patients carrying the T allele are less likely to resolve spontaneous HCV clearance (9). It is tempting to hypothesize that the lack of IFNλ4 production may also favor spontaneous clearance of SARS-CoV-2. In this regard, the frequency of the CC genotype was similar between the COVID-19 patients and a cohort of 309 patients with chronic hepatitis C in our hospital (37.85% vs. 34.60%, data not shown), suggesting a similar effect of IFNL4 polymorphisms in these diseases. However, it would be necessary to carry out further experiments such as analyzing these polymorphisms in asymptomatic individuals to confirm this hypothesis.

Our findings should be considered with caution, as the study has several limitations. The cohort of COVID-19 patients was not from a random selection, it was carried out in a single hospital, and we did not consider other potential confounding factors, such as comorbidities other than sex and age, which could influence the results. Therefore, our results should be interpreted as exploratory and descriptive until confirmed in further studies. However, some observations may support our findings. The allele and genotype frequencies in our control population without COVID-19 are comparable to those reported in similar populations (8,13). The polymorphism was not affected by sex, age, and severity of disease in COVID-19 patients as previously reported (14).

We propose that confirming our findings can help to understand some facts about COVID-19 that may be influenced by IFNL4 polymorphisms. In this regard, social and economic factors have been proposed to explain the disproportionate prevalence of COVID-19 in the African-descendant population in the USA (2,15). Our findings may provide an additional explanation on the basis that the T allele of rs12979860, which would be a risk factor for COVID-19 according to our results, is predominant in black populations (8,10,13). Additionally, treatments with type I IFNs have shown contradictory results in COVID-19 (4,5). It is tempting to speculate that, in addition to other factors (16), IFNL4 polymorphisms may affect the response to treatments with type I IFN as in chronic hepatitis C (10). Analysis of IFNL4 genetic variants may provide useful information to evaluate the efficacy of these treatments in COVID-19.

In summary, we have found that the IFNL4 rs12979860 polymorphism was associated with the presence of COVID-19 in a Spanish population. Confirmation of our results could have a substantial impact on the prognosis and treatment of this disease.

## Data Availability

No additional data but the included in the article.
The data that support the findings of this study are available from the corresponding author upon request

